# Effects of surgical masks on droplet and aerosol dispersion under various oxygen delivery modalities

**DOI:** 10.1101/2021.01.10.21249533

**Authors:** Takahiro Takazono, Kazuko Yamamoto, Ryuta Okamoto, Masato Tashiro, Shimpei Morimoto, Naoki Hosogaya, Taiga Miyazaki, Katsunori Yanagihara, Koichi Izumikawa, Hiroshi Mukae

**Author notes:** **Correspondence:** Takahiro TAKAZONO, M.D., Ph.D., Department of Infectious Diseases, Nagasaki University Graduate School of Biomedical Sciences, 1-7-1 Sakamoto, Nagasaki 852-8501, Japan. co-first author. **Author contributions:** T.T., K. Yamamoto, R.O., N.H., and H.M performed study design. T.T., K. Yamamoto, and R.O. supervised the experiments. T.T., K. Yamamoto, M.T., T.TM. K. Yanagihara, K.I., and H.M interpreted the data. S.M. supervised statical analysis. T.T. and K.Y. wrote the draft, and all the authors critically reviewed the manuscript and approved the final version of the manuscript. **All source(s) of support:** Financial support to conduct the study was provided by Fisher & Paykel Healthcare Co, Ltd. The funding source was not involved in the study design; in the collection, analysis, or interpretation of data; in the writing of the manuscript; or in the decision to submit the manuscript for publication.

## Abstract

**Rationale:** Aerosol dispersion under various oxygen delivery modalities, including high flow nasal cannula, is a critical concern for healthcare workers who treat acute hypoxemic respiratory failure during the coronavirus disease 2019 pandemic. Effects of surgical masks on droplet and aerosol dispersion under oxygen delivery modalities are not yet clarified.

**Objectives:** To visualize and quantify dispersion particles under various oxygen delivery modalities and examine the protective effect of surgical masks on particle dispersion.

**Methods:** Three and five healthy men were enrolled for video recording and quantification of particles, respectively. Various oxygen delivery modalities including high flow nasal cannula were used in this study. Particle dispersions during rest breathing, speaking, and coughing were recorded and automatically counted in each condition and were evaluated with or without surgical masks.

**Measurements and Main Results:** Coughing led to the maximum amount and distance of particle dispersion, regardless of modalities. Droplet dispersion was not visually increased by oxygen delivery modalities compared to breathing at room air. With surgical masks over the nasal cannula or high-flow nasal cannula, droplet dispersion was barely visible. Oxygen modalities did not increase the particle dispersion counts regardless of breathing pattens. Wearing surgical masks significantly decreased particle dispersion in all modalities while speaking and coughing, regardless of particle sizes, and reduction rates were approximately 95 and 80-90 % for larger (> 5 μm) and smaller (> 0.5 μm) particles, respectively.

**Conclusions:** Surgical mask over high flow nasal canula may be safely used for acute hypoxemic respiratory failure including coronavirus disease 2019 patients.

**Subject Category List:** 4.13 Ventilation: Non-Invasive/Long-Term/Weaning

*This article has an online data supplement, which is accessible from this issue’s table of content online at www.atsjournals.org.

## INTRODUCTION

Severe acute respiratory syndrome coronavirus-2 (SARS-CoV-2), which causes coronavirus disease 2019 (COVID-19), transmits by droplets and bioaerosols from COVID-19 patients (1). Based on the initial reports on the COVID-19 pandemic, around 5–30% of COVID-19 patients develop severe respiratory distress and require intensive care unit admission to receive advanced respiratory support in terms of oxygen therapy and non-invasive and invasive ventilatory support (2). High flow nasal cannula (HFNC) is a non-invasive oxygen delivery system that allows for administration of humidified air-oxygen blends as high as 60 L/min and a titratable fraction of inspired oxygen as high as 100%. Recent studies found that HFNC use in the management of respiratory failure caused by COVID-19 was associated with a reduced rate of invasive mechanical ventilation (3, 4). However, resources, including ventilators, as well as manpower of healthcare workers, during the COVID-19 pandemic are limited. Some patients with chronic hypoxemic respiratory insufficiency need palliative respiratory care, and some bedridden elderly patients do not wish to be intubated. Therefore, it is important to wisely utilize non-invasive respiratory care, including HFNC, for both patients and healthcare workers.

HFNC and non-invasive positive pressure ventilation (NPPV) have been categorized as aerosol-generating procedures (AGPs) (5) based on the hypothesis that the high-velocity gas flow may promote aerosolization of patients’ secretions containing viable viruses, which may then be dispersed in the environment and be inhaled by healthcare workers, thereby resulting in nosocomial infection during COVID-19 respiratory care (6). Therefore, evaluation of particle dispersion under these AGP-associated respiratory supports is critical for effective and safe treatment in severe COVID-19 patients. Some simulation studies showed that HFNC increased the dispersion amount and distance of large droplets (7, 8) and aerosols (9), while other clinical environmental studies found no significant increase in dispersed microbes (10) or aerosols (11) by HFNC. These findings indicate that the effects of HFNC on the dispersion of droplets and aerosols are controversial.

Notably, a study using coronavirus other than SARS-CoV-2 indicates that surgical masks reduce the number of droplets, aerosols, and viruses in the exhaled breath (12); however, no studies have investigated whether surgical masks prevent generation of droplets or small particles under respiratory management. Accordingly, this study aimed to visualize and quantify the particles generated from the human respiratory tract exposed to HFNC and other oxygen delivery modalities. Additionally, the effects of surgical masks on particle dispersion under oxygen delivery modalities were also investigated.

## METHODS

### Participants

Three and five healthy men (average height and weight: 173.6 (±7.4) cm and 68.6 (±5.6) kg, respectively) were voluntarily recruited for video recording and quantitative analysis of droplets, respectively. Inclusion criteria were as follows: 1) age ≥ 20 years; 2) no respiratory or systemic symptoms for 14 days before the experiments; and 3) no contact history with COVID-19 patients. Written informed consent was obtained from all the participants. All the staffs worked for these experiments wore the appropriate personal protective equipment including N95 mask and eye goggles inside the room. This study protocol was approved by the Nagasaki University Graduate School of Biomedical Sciences Research Ethics Committee (Approval number: 20092503).

### Visualization of droplet dispersion

In the visualization experiment, four different oxygen delivery modes, including nasal canula at 5 L/min (non-humidified, Nakamura Medical Industry Co., Ltd.), simple oxygen mask at 10 L/min (non-humidified, Nakamura Medical Industry Co., Ltd.), HFNC at 60 L/min (humidified, AIRVO2/Opti flow+, Fisher & Paykel Healthcare), and NPPV with an inspiratory positive airway pressure/expiratory positive airway pressure of 10/4 cm H_2_O at FiO_2_ of 21% (humidified, Philips Respironics V60/Amara Gel full face mask) were used. Particle dispersion under room air, nasal cannula, and HFNC and the dispersion patterns with or without surgical masks were recorded in the sitting position. Three breathing patterns, including rest breathing, 3 sets of speaking of sound (ta-chi-tsu-te-to and pa-pi-pu-pe-po), and 3 sets of coughing, were recorded in three participants. Generalized droplets from mouths were flashed continuously by Parallel Eye D (Shin Nippon Air Technologies). Scattering light from droplets was recorded by a super high-sensitive camera (Eye Scope, Shin Nippon Air Technologies) and analyzed by Image processing software (particle eye viewer, Shin Nippon Air Technologies). The recording area was 1 meter from participants’ mouths. The room temperature and humidity were maintained at 22 °C and 50%, respectively.

### Quantitative evaluation of droplet dispersion

For quantitative evaluation, the droplet dispersion under four conditions including room air, nasal canula at 5 L/min, and HFNC at 30 L/min or 60 L/min (humidified) and two patterns for each condition with or without surgical masks were measured. Also, the dispersion under three breathing patterns, including rest breathing for 30 seconds, 5 sets of speaking of sound (ta-chi-tsu-te-to and pa-pi-pu-pe-po), and 5 sets of coughing, were measured in five participants. Particles dispersed from participants’ mouths were recorded as described above and counted by the Fine Particle Visualization System (Type-S, Shin Nippon Air Technologies) with a 1/30 sec speed, which was located in two linear columns at 25-45 and 60-80 cm from the mouth, respectively (Supplement Figure 1). Particles sized > 5 µm and > 0.5 µm were automatically counted independently. The study was performed in an air-controlled room with a down-flow of 0.3 m/sec from the air supply system. The room temperature and humidity were maintained at 27 °C and 40%, respectively. Recorded movies were blinded to participants during both experiments, and participants had a 10 ml of water to keep the oral cavity hydrated between each experiment condition.

### Statistical Analysis

Differences in continuous numbers between the two groups were analyzed by ratio paired t-test. A p-value of < 0.05 was considered statistically significant. All the graphs were drawn using GraphPad Prism ver.8 (GraphPad Software).

## RESULTS

### Evaluation of visualization of droplet dispersion under various oxygen delivery modalities

Individual differences were observed in the amounts of droplets; however, three participants showed similar trends at each condition. The accumulated droplet (> 5 μm) dispersion in a representative participant is shown in Figure 1, and movies are shown in Supplemental videos. In the room air, droplet particles were barely visible at rest breathing but were observed while speaking and coughing. Some droplets reached over 60 and 100 cm from mouths while speaking and coughing. While coughing, visible particles had the largest amounts, the farthest distance, and the largest size. Visible droplet dispersion was neither increased nor farther reached among oxygen delivery modalities. Notably, by wearing a surgical mask over the nasal cannula or HFNC, droplet dispersion was barely visible, even while speaking and coughing.

**Figure 1.**
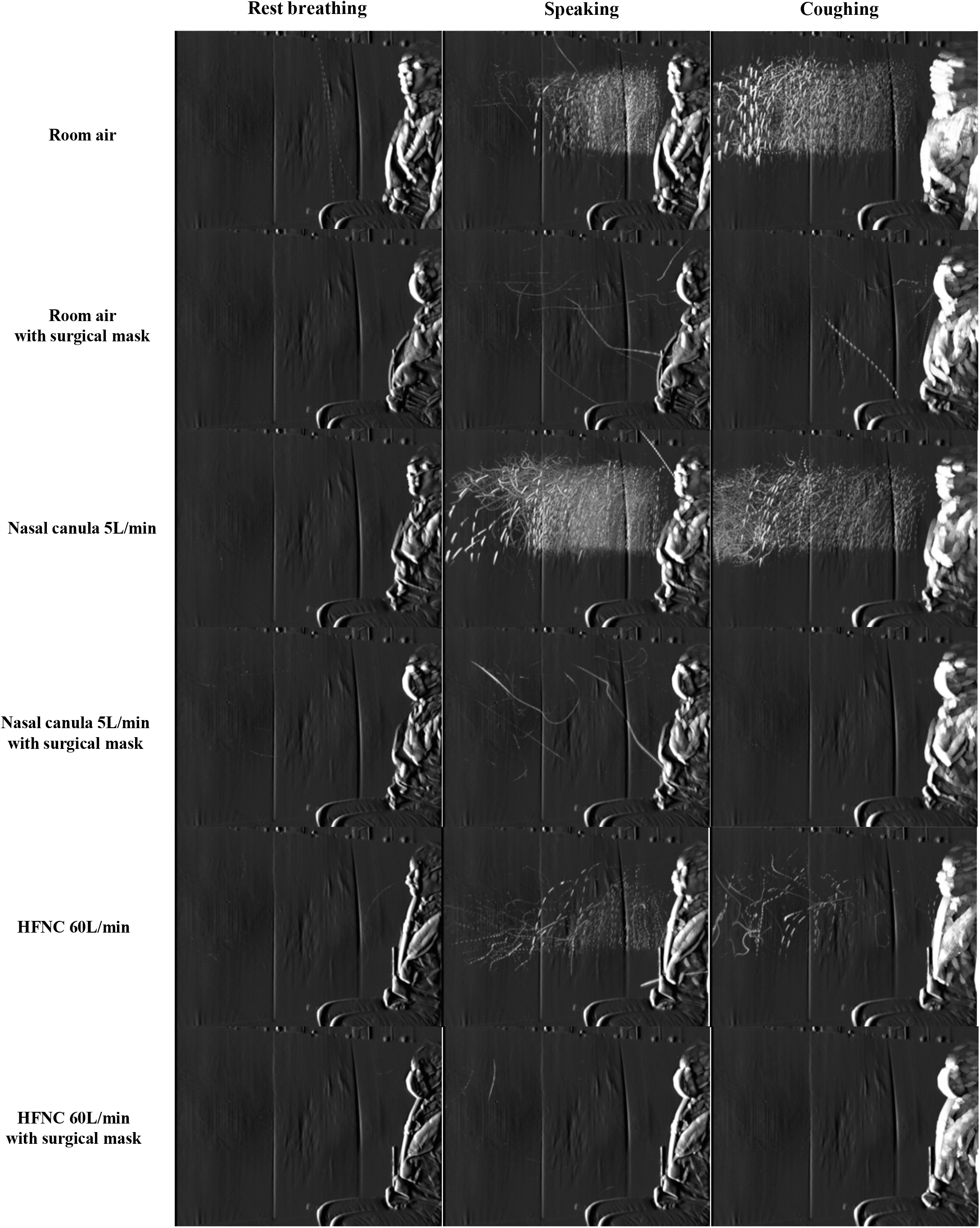
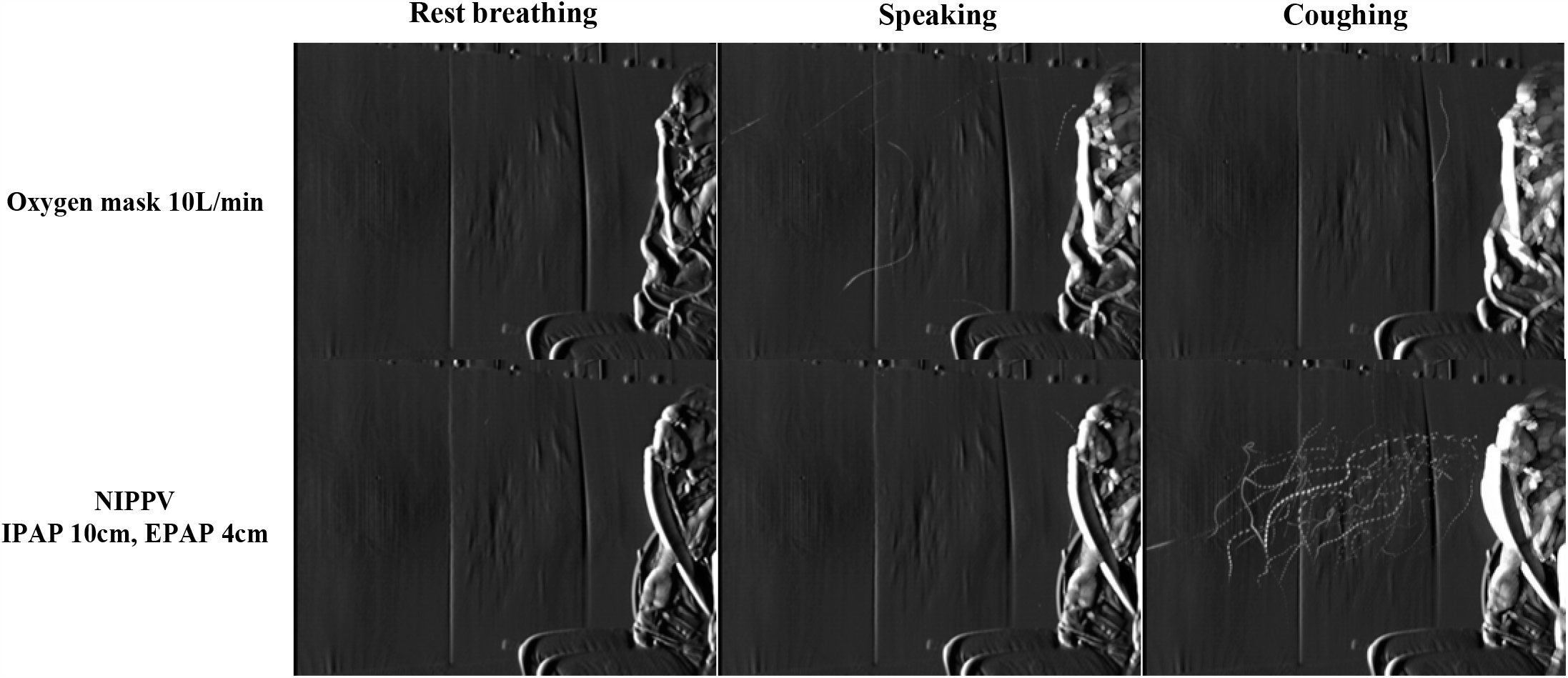
Representative accumulated photographs of droplet (> 5 µm) dispersion at rest breathing, speaking, and coughing at room air and with four different oxygen delivery modalities (5L/min of nasal cannula, 60L/min of HFNC, 10L/min of oxygen mask, and NIPPV(IPAP 10cm, EPAP 4cm)), with and without surgical masks. Abbreviations; HFNC, high flow nasal cannula.

With the simple oxygen mask of 10 L/min, small amounts of visible droplets floated from the slit between the oxygen mask and face while coughing. With the NPPV mode, droplets were visible through the expiration port of full-face NPPV masks and floated beyond the distance of 50 cm. These findings were observed in two of the three participants (Figure 1 and Supplement Figure 2 and 3).

### Quantitative evaluation of droplet dispersion under various oxygen delivery modalities

The quantification results of droplet and aerosol dispersion are shown in Figure 2. Droplet dispersion counts at coughing without surgical masks showed a 1-log increase compared to those at speaking and more than a 2-log increase compared to those at rest breathing. At rest breathing, particle counts were not different among all oxygen delivery modalities with surgical mask usage. Counts of droplets (> 5 μm) and smaller particles including aerosols (> 0.5 μm) were not different under nasal canula or HFNC and room air while speaking and coughing. Furthermore, the increased flow rate of HFNC (from 30 L/min to 60 L/min) did not affect the particle counts in each pattern, even while coughing.

**Figure 2.**
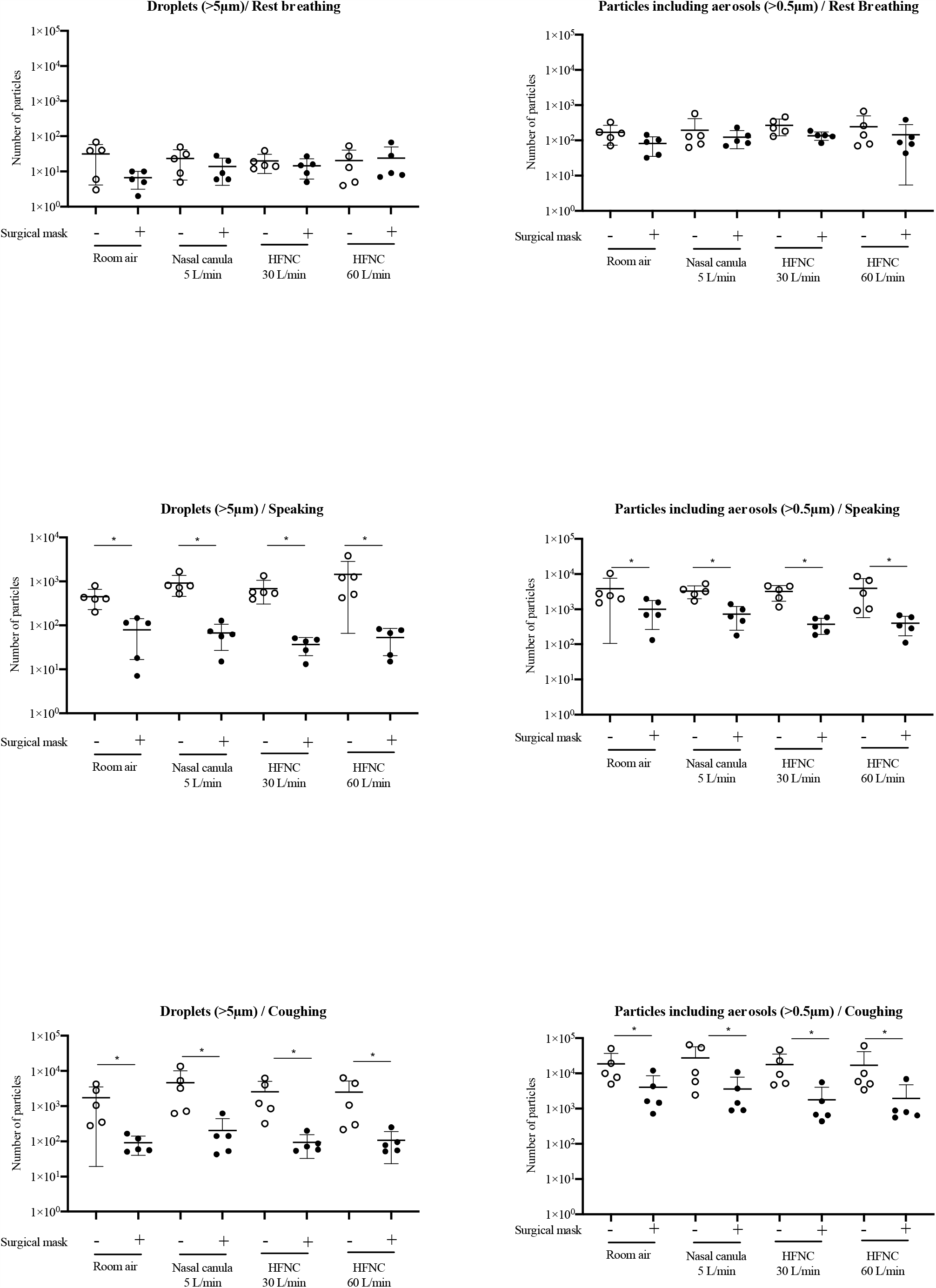
Number of droplet (>5 µm) and particles including aerosol (>0.5 µm) dispersion at room air and with three different oxygen delivery modalities (5L/min of nasal cannula, 30L/min or 60L/min of HFNC), in three breathing patterns (rest breathing, speaking, and coughing), with or without surgical masks. Abbreviations; HFNC, high flow nasal cannula. * *P* < 0.05, analyzed by ratio paired t-test.

We examined the reduction ratios of wearing surgical masks in the particle count of each size under each oxygen delivery modality at each breathing pattern (Table 1). As a result, at speaking and coughing under the nasal cannula or the HFNC, we observed decreases in the particle counts by wearing surgical masks (P < 0.05, the t-test for the null-difference within a participant between the usage of the surgical mask.). At speaking and coughing, reduction ratios of particle dispersion by wearing surgical masks were approximately 95% and 80-90% in droplets (> 5 μm) and smaller particles including aerosols (> 0.5 μm), respectively (Table 1). Interestingly, reduction efficacy of dispersion by surgical masks tended to be higher under NHFC than under nasal cannula while speaking.

**Table 1.** Reduction effect of wearing surgical masks on particle dispersion with various oxygen delivery modalities.

| Reduction ratio (Surgical mask +/- ) |  |  |  |  |
| --- | --- | --- | --- | --- |
|  |  | Average (min - max) % |  |  |
| Particle sizes | Oxygen delivery modes | Rest breathing | Speaking | Coughing |
| >5 µm | Room air | 21.15 (5 - 333.3) | 17.81 (3.5 - 33.7) | 5.15 (2 - 21.4) |
|  | Nasal canula (5 L/min ) | 58.97 (19.4 -120) | 7.39 (1.9 - 17.6) | 4.28 (0.8 - 19.7) |
|  | HFNC (30 L/min) | 72.73 (23.1 - 180) | 5.35 (1 - 10.9) | 3.70 (1.3 - 18.1) |
|  | HFNC (60 L/min) | 116.67 (61.5 - 255) | 3.64 (0.6 -15.3) | 4.29 (2.2 - 26.3) |
| >0.5 µm | Room air | 48.0 (24.7 - 119.2) | 26.39 (8.7 - 64.6) | 21.71 (14.3 - 23.8) |
|  | Nasal canula (5L/min ) | 62.79 (39.9 - 121.5) | 22.03 (8.8 - 38) | 13.01 (5.8 - 37.1) |
|  | HFNC (30 L/min) | 50.45 (27.9 - 106.9) | 11.65 (5.1 - 25.5) | 10.11 (4.6 - 14.2) |
|  | HFNC (60 L/min) | 59.03 (30.4 - 157) | 10.09 (1.3 - 37.4) | 11.50 (8.7 - 16.3) |

In terms of dispersion distance of droplets and aerosols, the number of particles obtained from two separate counters is shown in Figure 3. Dispersion distance of droplets (> 5 μm) and smaller particles including aerosols (> 0.5 μm) decreased at the far side compared to the near side. Smaller particles including aerosols (> 0.5 μm) obviously decreased at the far side compared to the near side while speaking; however, they remained at the far side while coughing. Surgical masks effectively reduced aerosol dispersion, even while coughing. Compared to nasal cannula, HFNC did not increase aerosol dispersion in the near or far side. Moreover, the flow rate of 60 L/min did not increase aerosol dispersion compared to that of 30 L/min in the near or far side.

**Figure 3.**
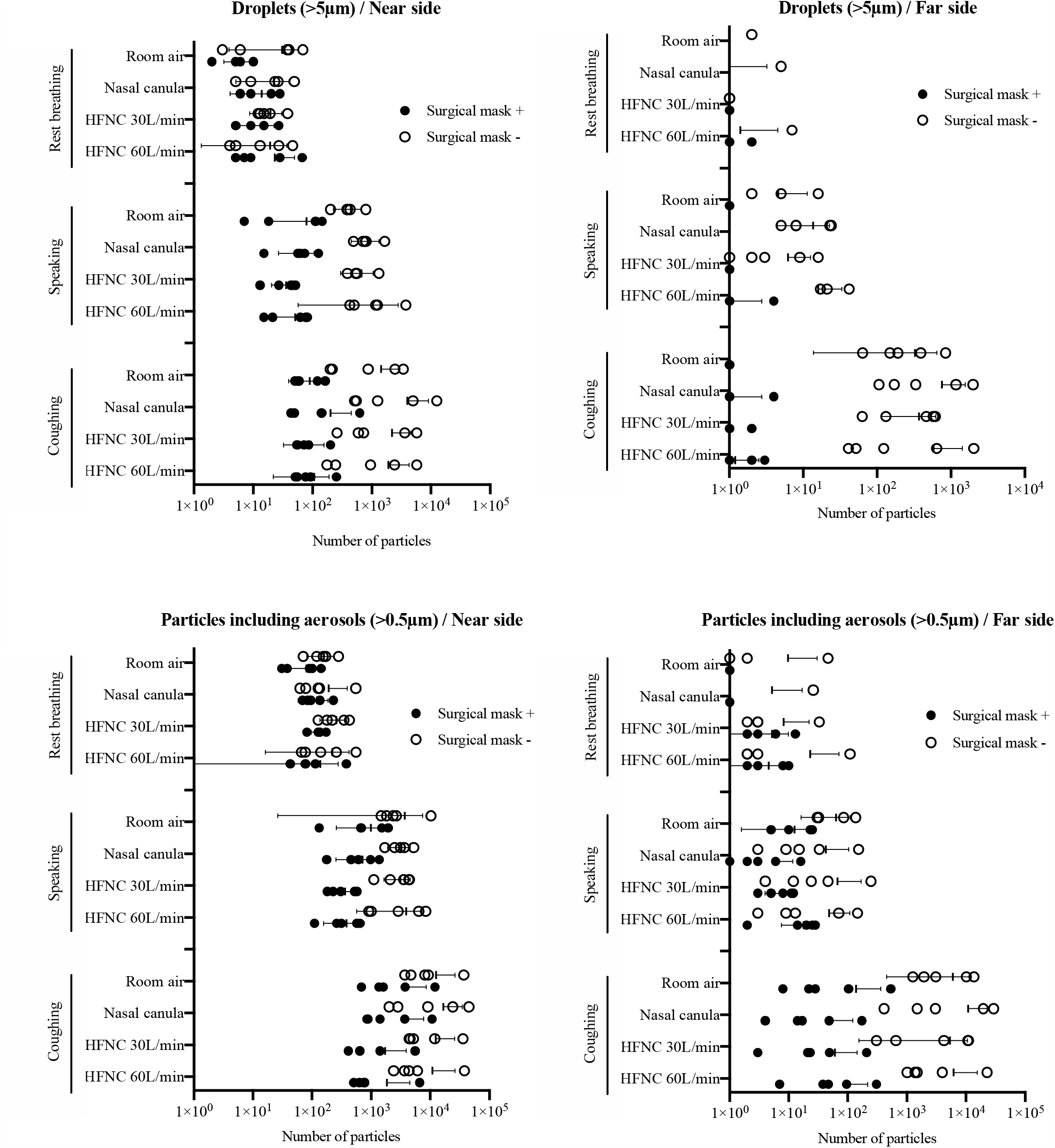
Numbers of droplet (>5 µm) and particles including aerosol (>0.5 µm) dispersion and their distance obtained by two separate counters (near side: 25-45 cm and far side: 60-80 cm from the mouth). Those particles were counted at room air and three different oxygen delivery modalities (5L/min of nasal cannula, 30L/min or 60L/min of HFNC), in threebreathing patterns (rest breathing, speaking, and coughing), with or without surgical masks. Abbreviations; HFNC, high flow nasal cannula.

## DISCUSSION

In this study, we performed two experiments in different oxygen delivery modes and modalities. Based on the particle visualization study, we found that visible droplets reached 60–80 cm while speaking and 1 m while coughing in a sitting position; the findings are similar to those in a recent study (13). In addition, these findings support results in a systematic meta-analysis, which showed that a physical distance of 1 m or more is optimum to reduce person-to-person virus transmission and to keep healthcare workers from contracting the SARS-CoV-2 infection (14). Surgical masks obviously reduced visible droplets in our study; these findings in photos and videos confirm the effect of surgical masks on transmission of viral particles in society and in hospitals caring for oxygen-delivered patients during the pandemic. Based on our quantification study, the majority of particles produced by the HFNC mode were smaller particles or aerosols; the finding is supported by a previous study, which measured the median diameter of particles (range, 1.19-1.42 μm) produced by HFNC (11). Coughing increased the amount and distance of dispersing smaller particles, suggesting that aerosols from lung alveoli were especially dispersed by coughing.

HFNC did not increase the number of smaller particles or aerosols compared to conventional nasal cannula or room air while coughing. These findings suggest that oxygen delivery devices investigated in this study are potential aerosol-dispersing procedures, but not AGPs (15).

In the NPPV mode, although only one type of NPPV mask was evaluated in this study, floated droplets were visible while coughing and might harm bioaerosol dispersion during treatment. In addition, droplets and aerosols were approximately 10-times more during coughing than during speaking, regardless of types of oxygen devices. The difference is larger than that in a previous study (11), which showed about a twice increased number of particles while coughing in any devices; however, those particles were assessed only about 5cm away from mouths in that study (11). Notably, surgical masks significantly reduced dispersion counts of both droplets and aerosols while speaking or coughing, regardless of oxygen delivery modes. Indeed, surgical masks effectively reduced over 90% of droplets and over 95% of aerosols during HFNC. While speaking, the reduction rate of particle dispersion by surgical masks during HFNC was higher than that during nasal cannula. This result is supported by previous findings of velocity map of gas flow experiments (16), which showed that the proportion of droplets (> 5 µm) captured by surgical masks was higher during 40 L/min of HFNC than during 6 L/min of nasal cannula, suggesting that high-velocity particles are more likely to be captured into surgical masks than lower velocity particles (16). In our study, 60 L/min of HFNC did not increase counts and dispersion distance of droplets or aerosols compared to 30 L/min of HFNC; the finding is consistent with that in a previous study (11). The particle dispersion may be limited when the cannula of HFNC is properly fitted (9), even at the highest flow rate.

This study has several limitations. First, this study included a relatively small number of individuals. The particle experiments were performed in healthy volunteers, but not in viral pneumonia patients. Second, only one-sided visualization was recorded, and dispersion in the three-dimensional space was not evaluated. Third, the particle quantification experiment was performed at a mild down flow, which might affect the natural distance of dispersion, and the design of the quantification experiment limited the detection area of particles. Forth, particles with a size of less than 0.5 µm were not detectable in our study; however, these smaller particles are not speculated to contain enough viruses for transmission (17). To date, the virus load of SARS-CoV-2 in aerosols generated from coughing or included in exhaled breath from patients with COVID-19 has not been reported (18), although SARS-CoV-2 has been found to be viable in room air for 3 hours (19). The virus load or concentration required for transmission in humans needs to be further investigated.

In summary, with our study, HFNC did not increase either quantity or distance of droplet and aerosol dispersion compared to standard nasal cannula therapy, and surgical masks decreased particle dispersion and improved safety of healthcare workers during HFNC, even at the highest flow, in appropriate infection control settings. Our findings support that surgical masks should be recommended for patients with acute hypoxemic respiratory failure due to viral pneumonia, including COVID-19 patients. Further studies are warranted to examine viral spread and risks associated with HFNC therapy in COVID-19 patients.

## Data Availability

The data are not publicly available as containing information could compromise the privacy of research participants

## Acknowledgement

We would like to thank technical staffs in Shin Nippon Air Technologies for their supports in this study.

## ONLINE DATA SUPPLEMENT

### Supplemental videos

The movie of droplet (> 5 *μ* m) dispersion in a representative participant at rest breathing, while speaking, and while coughing at room air and with four different oxygen delivery modalities, including a nasal canula at 5 L/min, a simple oxygen mask at 10 L/min, HFNC at

60 L/min, and NPPV at an inspiratory positive airway pressure/expiratory positive airway pressure of 10/4 cm H_2_O at FiO_2_ of 21% with or without surgical mask. Abbreviations; HFNC, high flow nasal cannula.

**Supplemental Figure 1**.

Schematic diagram of methods for quantitative evaluation of droplets production. Dispersing particles sized > 5 µm and > 0.5 µm were counted by the Fine Particle Visualization System (Type-S, Shin Nippon Air Technologies), which was located in two linear columns at 25-45 and 60-80 cm from the mouth, respectively.

**Supplemental Figures 2 and 3** Accumulated photographs of droplet (> 5μm) dispersion with two other participants. At rest, while breathing, while speaking, and while coughing at roomair and with three different oxygen delivery modalities, including a nasal canula at 5 L/min, HFNC at 30 L/min, and at 60L/min with or without surgical mask. Abbreviations; HFNC, high flow nasal cannula.

