## Supplementary material for "Effects of surgical masks on droplet and aerosol dispersion under various oxygen delivery modalities": Supplemmental Figure 1, 2, and 3

### Slide 1
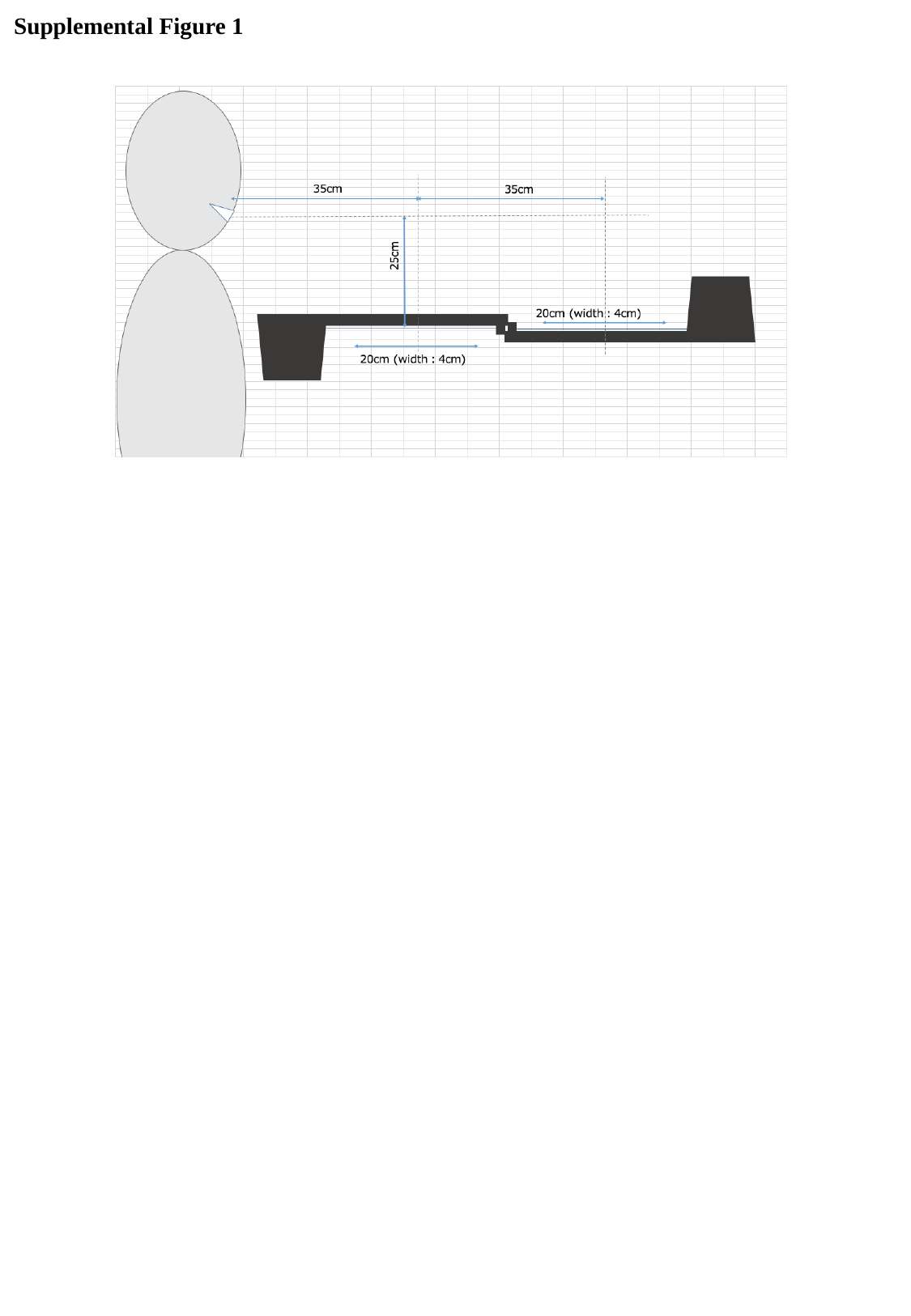

Supplemental Figure 1

### Slide 2
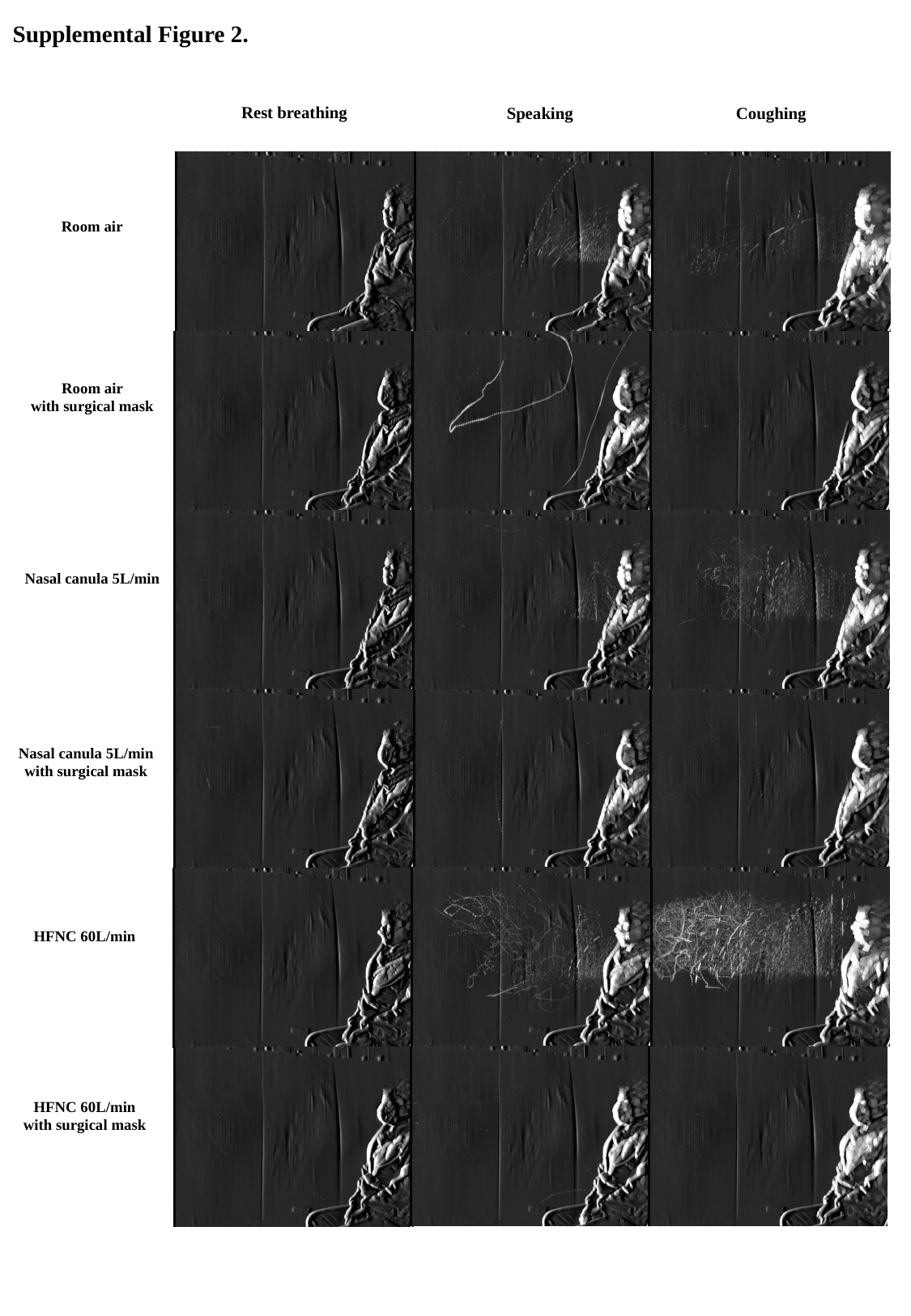

Supplemental Figure 2.
Rest breathing
Speaking
Coughing
Room air
Room air
with surgical mask
Nasal canula 5L/min
Nasal canula 5L/min
with surgical mask
HFNC 60L/min
HFNC 60L/min
with surgical mask

### Slide 3
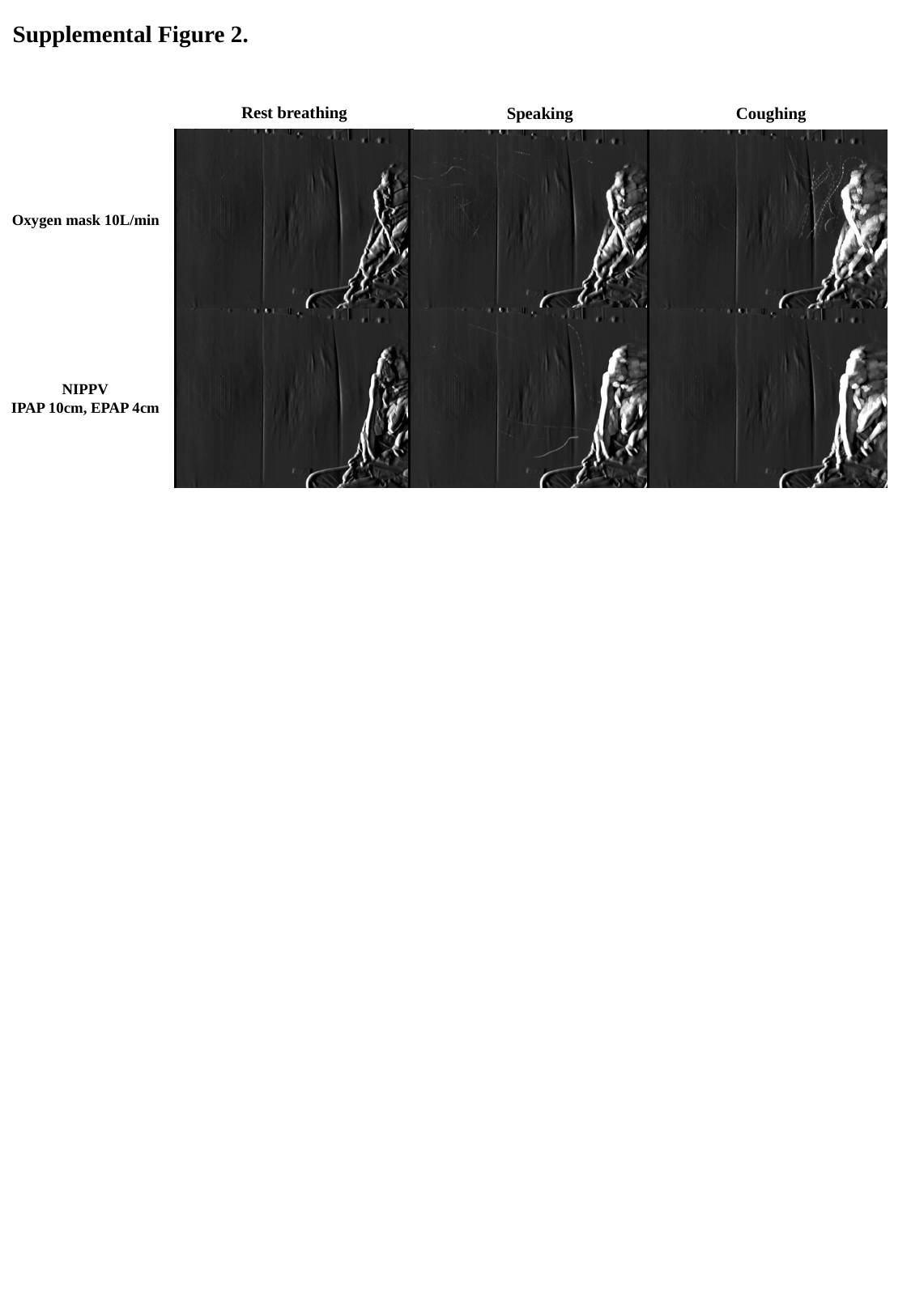

Supplemental Figure 2.
Rest breathing
Speaking
Coughing
Oxygen mask 10L/min
NIPPV
IPAP 10cm, EPAP 4cm

### Slide 4
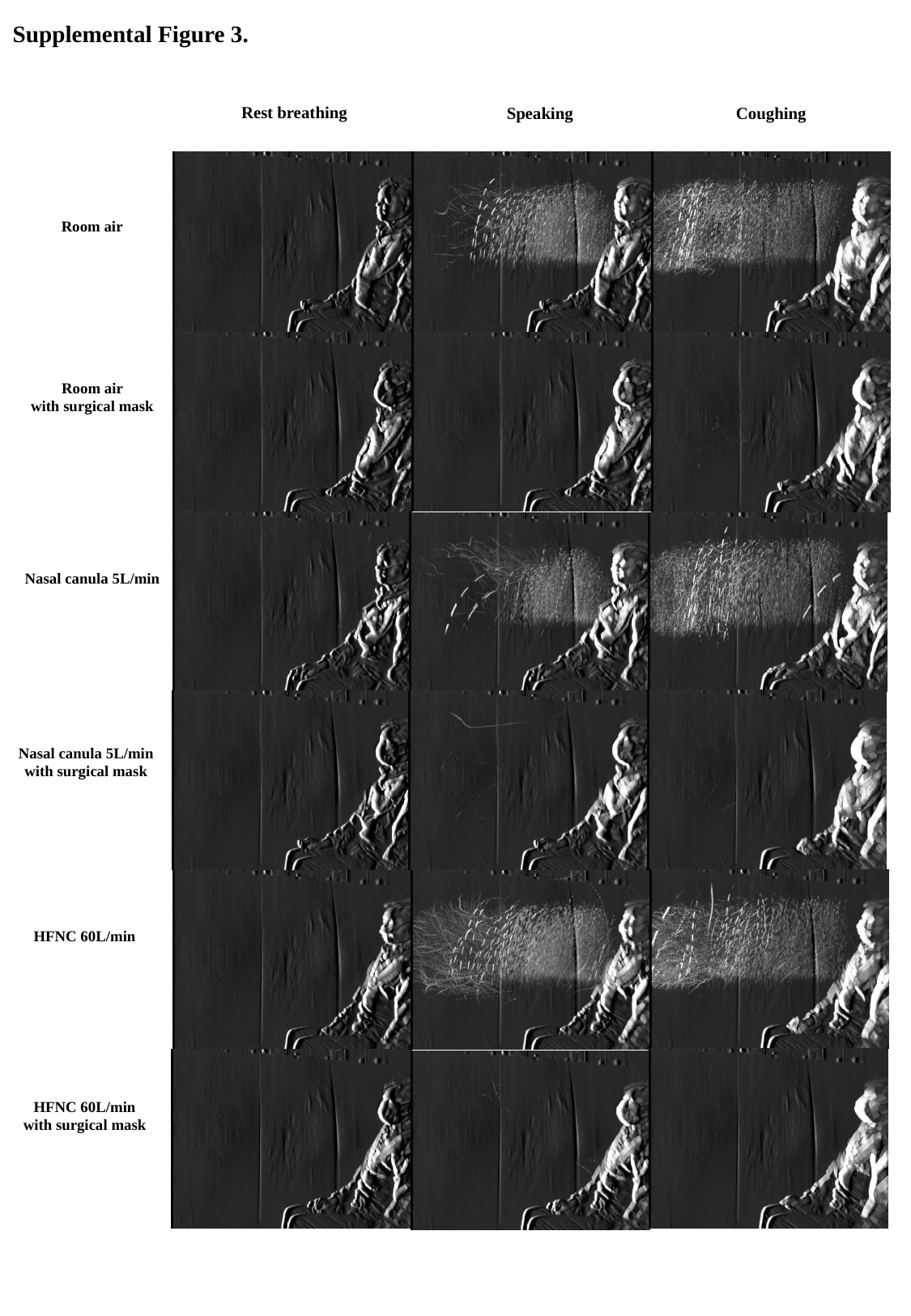

Supplemental Figure 3.
Rest breathing
Speaking
Coughing
Room air
Room air
with surgical mask
Nasal canula 5L/min
Nasal canula 5L/min
with surgical mask
HFNC 60L/min
HFNC 60L/min
with surgical mask

### Slide 5
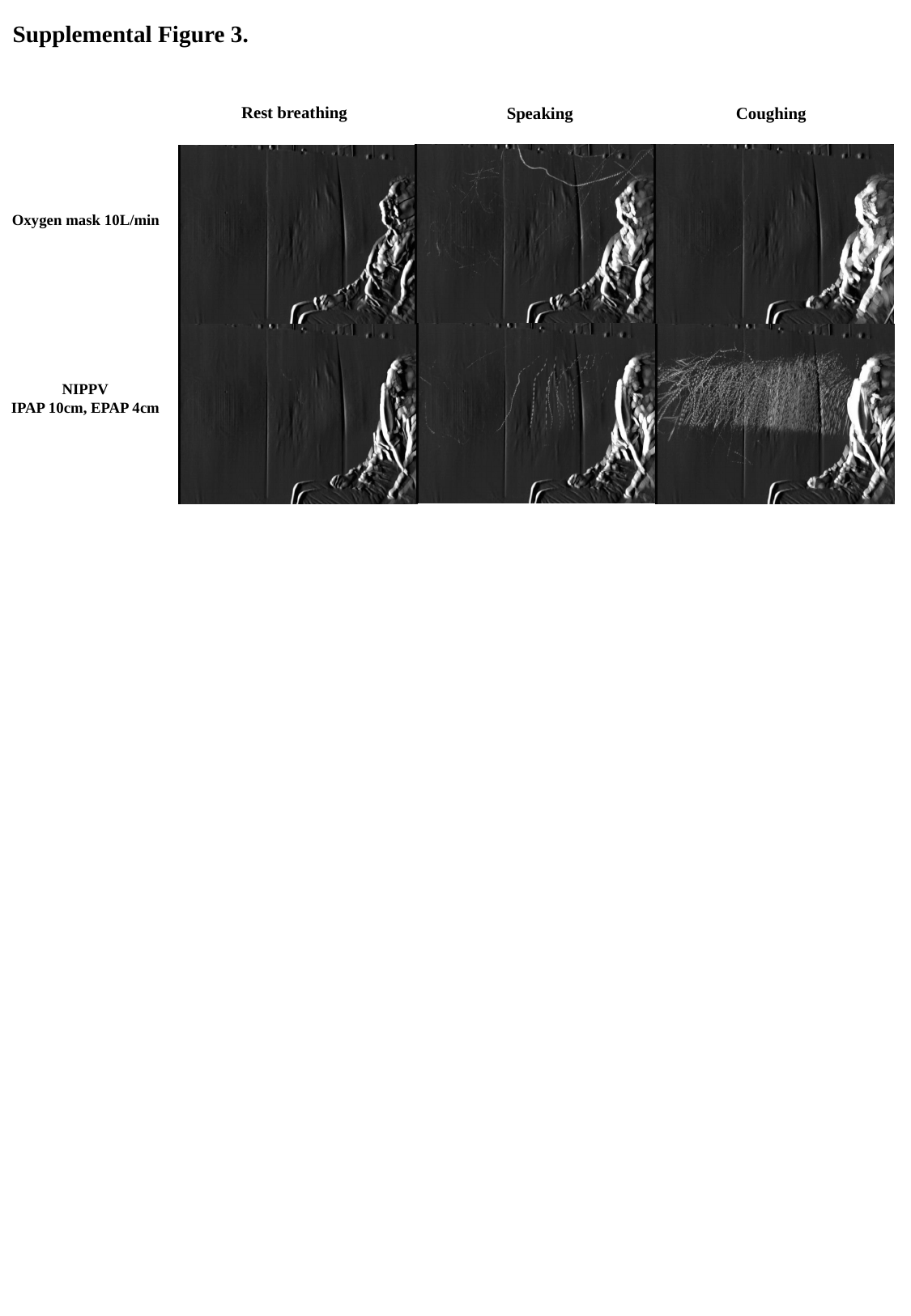

Supplemental Figure 3.
Rest breathing
Speaking
Coughing
Oxygen mask 10L/min
NIPPV
IPAP 10cm, EPAP 4cm
